# Mpox (monkeypox) knowledge, concern, willingness to change behaviour, and seek vaccination: Results of a national cross-sectional survey

**DOI:** 10.1101/2022.12.01.22282999

**Authors:** James MacGibbon, Vincent Cornelisse, Anthony K J Smith, Timothy R Broady, Mohamed A Hammoud, Benjamin R Bavinton, Heath Paynter, Matthew Vaughan, Edwina J Wright, Martin Holt

## Abstract

**Background:** We assessed knowledge and concern about mpox, acceptability of behavioural changes to reduce transmission risk, and willingness to be vaccinated among gay, bisexual and queer-identifying men and non-binary people.

**Methods:** We conducted a national, online cross-sectional survey with a convenience sample between August and September 2022. Participants were recruited through community organisation promotions, online advertising, and direct email invitations. Eligible participants were gay, bisexual or queer; identified as male (cisgender or transgender) or non-binary; aged 16 years or older; and lived in Australia. The main outcome measures were: knowledge and concern about mpox; recognition of mpox symptoms and transmission routes; vaccination history; acceptability of behavioural changes to reduce mpox risk, and willingness to be vaccinated.

**Results:** Of 2287 participants, most participants were male (2189/2287; 95.71%) and gay (1894/2287; 82.82%). Nearly all had heard about mpox (2255/2287; 98.60%), and the majority were concerned about acquiring it (1461/2287; 64.42%). Most of the 2268 participants not previously diagnosed with mpox identified skin lesions (2087; 92.02%), rash (1977; 87.17%), and fever (1647; 72.62%) as potential symptoms, and prolonged and brief skin-to-skin contact as potential ways to acquire mpox (2124, 93.65%; and 1860, 82.01% respectively). The most acceptable behavioural changes were reducing or avoiding attendance at sex parties (1494; 65.87%) and sex-on-premises venues (1503; 66.40%), and having fewer sexual partners (1466; 64.64%). Most unvaccinated and undiagnosed participants were willing to be vaccinated (1457/1733; 84.07%).

**Conclusions:** People at risk of mpox should be supported to adopt acceptable risk reduction strategies during outbreaks and seek vaccination.

## Introduction

Mpox (formerly ‘monkeypox’) is a viral zoonotic disease first identified in humans in 1970 and is endemic in Central and West African countries (1). In May 2022, health authorities identified an emerging mpox outbreak in non-endemic countries, in which >95% cases were recorded among gay and bisexual and other men who have sex with men (GBMSM) (1, 2). In July 2022, the World Health Organization declared mpox a public health emergency of international concern (2). Australia made mpox a nationally notifiable disease in June 2022 and declared the mpox outbreak a Communicable Disease Incident of National Significance (CDINS) in July 2022 (3). In late November 2022, the CDINS declaration was withdrawn (3).

Mpox spreads primarily through direct contact with infected skin and bodily fluids, and by large respiratory droplets (4). Despite initial concerns of fomite transmission via contaminated surfaces, to date no such transmission has been documented during the current outbreak (4, 5). Mpox infection is self-limited and requires self-isolation until lesions have healed (6, 7). Clinical care, including hospitalisation may be required to manage secondary bacterial infections, ocular and gastrointestinal involvement and severe pain (6, 7). Mpox is rarely fatal (6, 7). More than 85,000 mpox cases have been recorded in 103 non-endemic countries, with 81 fatalities (8). Globally, the number of incident infections peaked in August 2022 at around 1,000 cases daily, and have declined since then (9). Australia recorded its first mpox cases in May 2022, and 144 cases by March 2023, two-thirds of which were acquired overseas (8).

Vaccinia vaccines, developed to protect against smallpox, offer protection against mpox (10, 11). In August 2022, the Australian Government acquired a limited supply of 3^rd-^generation, non-replicating Modified Vaccinia Ankara (MVA) vaccine, initially targeted to people at highest risk of exposure to monkeypox virus or severe mpox illness (12, 13). As was observed during COVID-19, particularly before widespread vaccine availability (14, 15), GBMSM will likely follow public health advice, promoted via community organisations, and adapt their behaviour to reduce their risk of acquiring monkeypox virus until they are vaccinated; for example by reducing the number of untraceable sexual partners and attendance at sex-on-premises venues and sex parties. Based on experience with COVID-19, we anticipate most GBMSM will be willing to be vaccinated against mpox (16, 17).

To identify education and health promotion needs for people at risk of mpox, and help guide the Australian vaccination program, we examined what GBMSM and non-binary people knew about monkeypox virus and what behavioural changes they would be prepared to undertake in response to the outbreak. We anticipated that greater concern about mpox would be associated with a greater willingness to modify behaviour and to seek vaccination.

## Method

### Study design and participants

A national Australian, cross-sectional survey was conducted between 24 August and 12 September 2022 using Qualtrics software (Provo, UT). The survey was promoted through community partner organisations and paid advertisements on Facebook and Grindr, supplemented by email invitations to consenting participants of previous studies. Potential participants were directed to the study website, containing participant information and the survey link. Participants provided consent before starting the survey. Eligible participants identified as gay, bisexual or queer (GBQ); identified as male (cisgender or transgender) or non-binary; were aged 16 years or older; and lived in Australia. The study was approved by the ethics committee of UNSW Sydney (HC220484) and the community organisation ACON (2022/14).

### Measures

We collected data on demographics, physical health, recent sexual practices, HIV status, HIV treatment or pre-exposure prophylaxis (PrEP), sexually transmissible infection (STI) testing and history, vaccination history against smallpox or mpox, knowledge and concern about mpox, acceptability of behavioural changes in response to mpox, and willingness to be vaccinated. Adaptive routing was used to exclude irrelevant items, and response options were randomised in lists to reduce order bias. Participants who had been diagnosed with mpox were shown questions about their experience of mpox (not discussed in this article). The main outcome measures are detailed in Appendix 1 (see supplementary materials).

### Statistical analyses

Descriptive statistics were computed for all variables. Pearson’s chi-squared tests identified significant associations between independent and dependent variables, and logistic regression was used to identify independent associations for key dependent variables (i.e., behavioural change measures and vaccine willingness). Multivariable analyses used block entry of variables that were significant at the bivariable level. Statistical assumptions were assessed, including model diagnostics for logistic regression, none of which were violated. Variables in the regression models had no missing observations. We report unadjusted and adjusted odds ratios (OR and aOR) with 95% confidence intervals (CI). Statistical significance was set at *p*<.05 (two-tailed). Analyses were conducted using Stata version 16.1 (StataCorp, College Station, TX).

## Results

The survey was completed by 2287 of 3232 eligible people who commenced it (70.76% completion rate). The median age of the 2287 participants was 40 years (IQR=31–51), 1894 (82.82%) identified as gay, 2189 (95.71%) were male, 1701 (74.38%) were Australian born, 1550 (67.77%) were university educated, 1647 (72.02%) reported full-time employment, and 1877 (82.07%) lived in the capital city of their state/territory. Most participants lived in New South Wales (860; 37.60%) or Victoria (760; 33.20%). In total, 1944 (85.00%) were HIV-negative, 179 (7.83%) were HIV-positive, and 164 (7.17%) were untested or did not know their status. Within the prior 12 months, more than two thirds of the sample had been tested for HIV (1634; 71.45%) or other STIs (1616; 70.66%). In total, 1241 (54.27%) participants had ever used PrEP and 914 (39.97%) were taking PrEP at the time of the survey. Further details of participant characteristics are shown at Appendix 2, Table 1 (see supplementary materials).

### Previous vaccination against smallpox or mpox

Nearly one quarter of participants had ever received a smallpox or mpox vaccine (541/2287; 23.66%). Most had received MVA vaccine (325/541; 60.1% of vaccinated participants or 14.21% of the total sample), a small proportion had received replication-competent 2^nd^-generation vaccinia vaccine (ACAM2000) (28/541; 5.2%), and more than one third did not know which vaccine they had received (188/541; 34.8%). Most were vaccinated after May 2022 (347/541; 64.1%), and had received the vaccine before potential exposure to mpox (283/347; 81.6%). Fifteen participants were vaccinated after potential exposure (15/347; 4.3%), and the remainder reported they received the vaccine for ‘another reason’; that is, unrelated to potential exposure to mpox (49/357; 14.1%). Of participants vaccinated with MVA, most had received only one dose (320/325; 98.5%) and most received it in Australia (295/325; 90.8%). Further details of previous vaccination are shown at Appendix 2, Table 2.

### Potential mpox risk factors

Of the 2287 participants, 706 (30.87%) had travelled overseas in the previous six months, and 1075 (47.00%) planned to travel in the next six months. In the six months preceding the survey, 599 (26.19%) participants had more than 10 male sexual partners, 1277 (55.84%) reported casual sex with male partners without condoms, 1248 (54.58%) reported anonymous sex, 804 (35.16%) reported group sex, and 731 (31.96%) had visited a sex-on-premises venue. Compared with unvaccinated participants, participants vaccinated since May 2022 were more likely to report recent and future overseas travel, and other potential mpox risk factors (see Appendix 2, Table 3 for comparisons).

### Mpox knowledge and concern

Of the 2268 participants who had not been diagnosed with mpox, 32 (1.41%) had never heard of mpox before the survey, 1065 (46.96%) knew a ‘small amount’, 695 (30.65%) a ‘fair amount’, 356 (15.70%) ‘quite a bit’ and 120 (5.29%) ‘a lot’. The most common ways to learn about mpox were the media (1904/2255; 84.43%), conversations with friends or family (729/2255; 32.33%), and information provided by community organisations (660/2255; 29.27%). Among the 2268 undiagnosed participants, 323 (14.24%) knew someone who had been diagnosed with mpox, 1461 (64.42%) were concerned or very concerned about acquiring mpox, and 465 (20.50%) believed it likely or very likely they would acquire mpox. Vaccinated participants were more likely to report mpox knowledge and concern compared to unvaccinated participants, but there was no difference in perceived likelihood of acquiring mpox (see Appendix 2, Table 4 for all comparisons).

### Recognition of mpox symptoms and transmission routes

The 2268 undiagnosed participants were asked to identify potential mpox symptoms and transmission routes. The most commonly identified symptoms were skin lesions (2087; 92.02%), skin rash (1977; 87.17%), and fever (1647; 72.62%). Fig. 1 shows other symptoms and uncertainty about potential symptoms (ranging from 5–43% for ‘don’t know’ responses).

**Figure 1.**
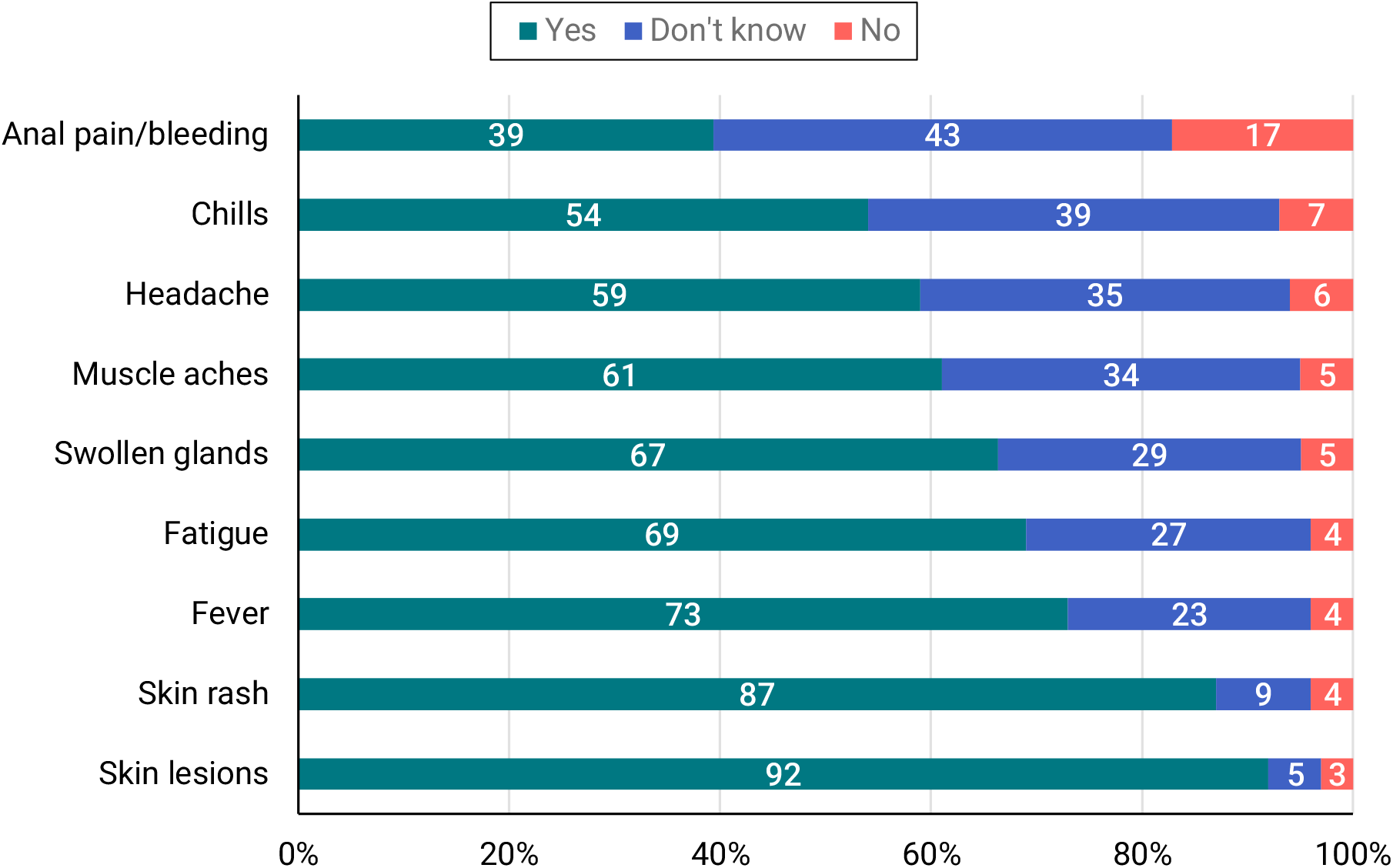
– Mpox symptom identification by 2268 participants who had not been diagnosed with mpox. All potential mpox symptoms were presented in random order. Three ‘decoy’ symptoms were included in the survey but are not reported here, e.g., ‘loss of taste or smell’.

Most participants identified prolonged skin-to-skin contact (2124; 93.65%), brief skin-to-skin contact (1860; 82.01%) and contact with bodily fluids (1692; 74.60%) as potential ways to acquire mpox. Fig. 2 shows knowledge of transmission routes and uncertainty about them (ranging from 5–36%). Participants with more than 10 male sexual partners in the previous six months were more likely to recognise potential mpox symptoms (and some transmission routes) compared to those with fewer partners (see Appendix 2, Table 5 for all comparisons).

**Figure 2.**
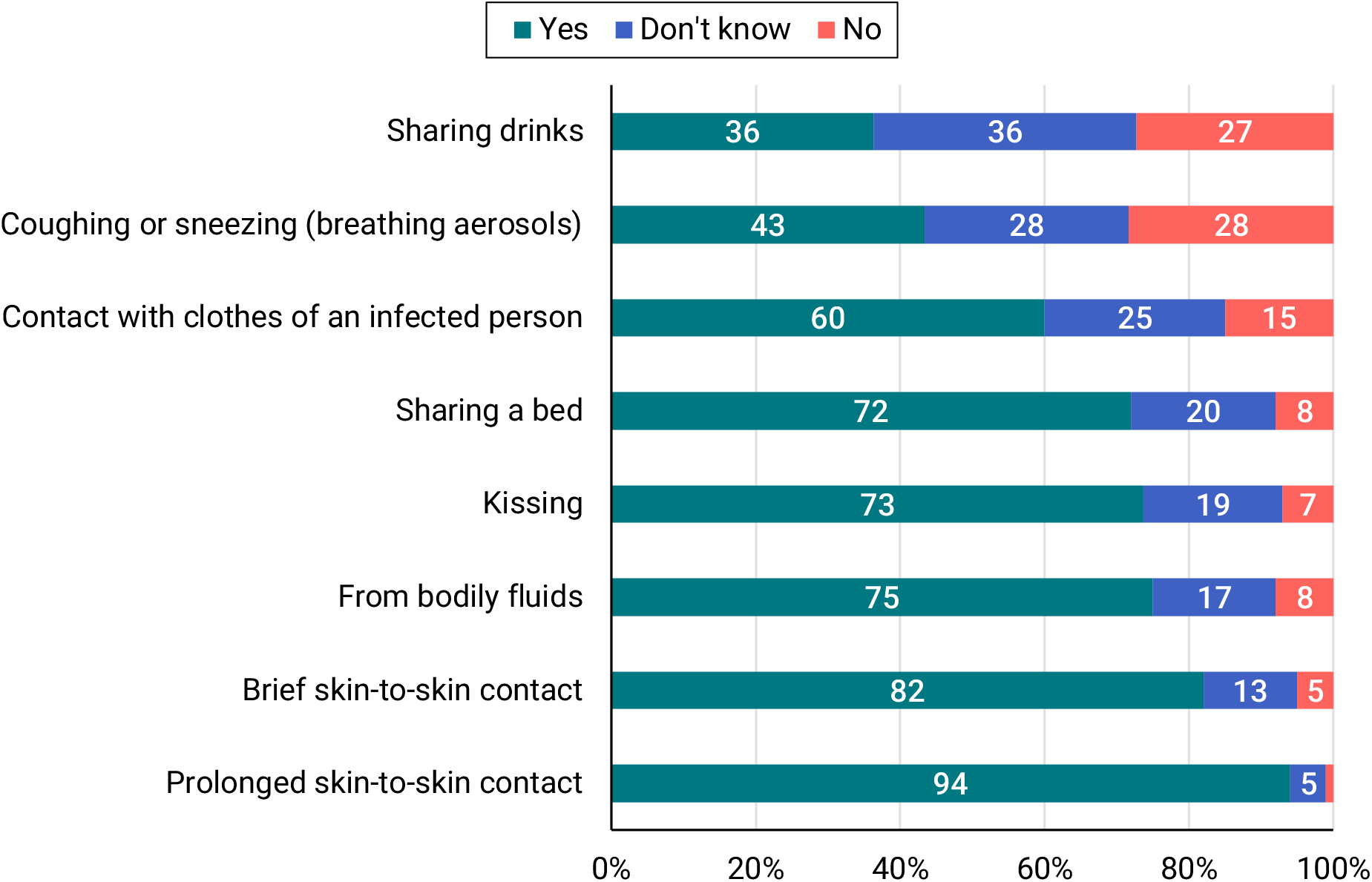
– Mpox transmission route identification by 2268 participants who had not been diagnosed with mpox. All potential mpox transmission routes were presented in random order.

### Acceptability of behavioural changes in response to mpox

Fig. 3 shows the acceptability of different behavioural changes among the 2268 undiagnosed participants. The most acceptable strategies were reducing or avoiding attendance at sex parties (1494; 65.87%) and sex-on-premises venues (1503; 66.40%), and having fewer sexual partners (1466; 64.64%).

**Figure 3.**
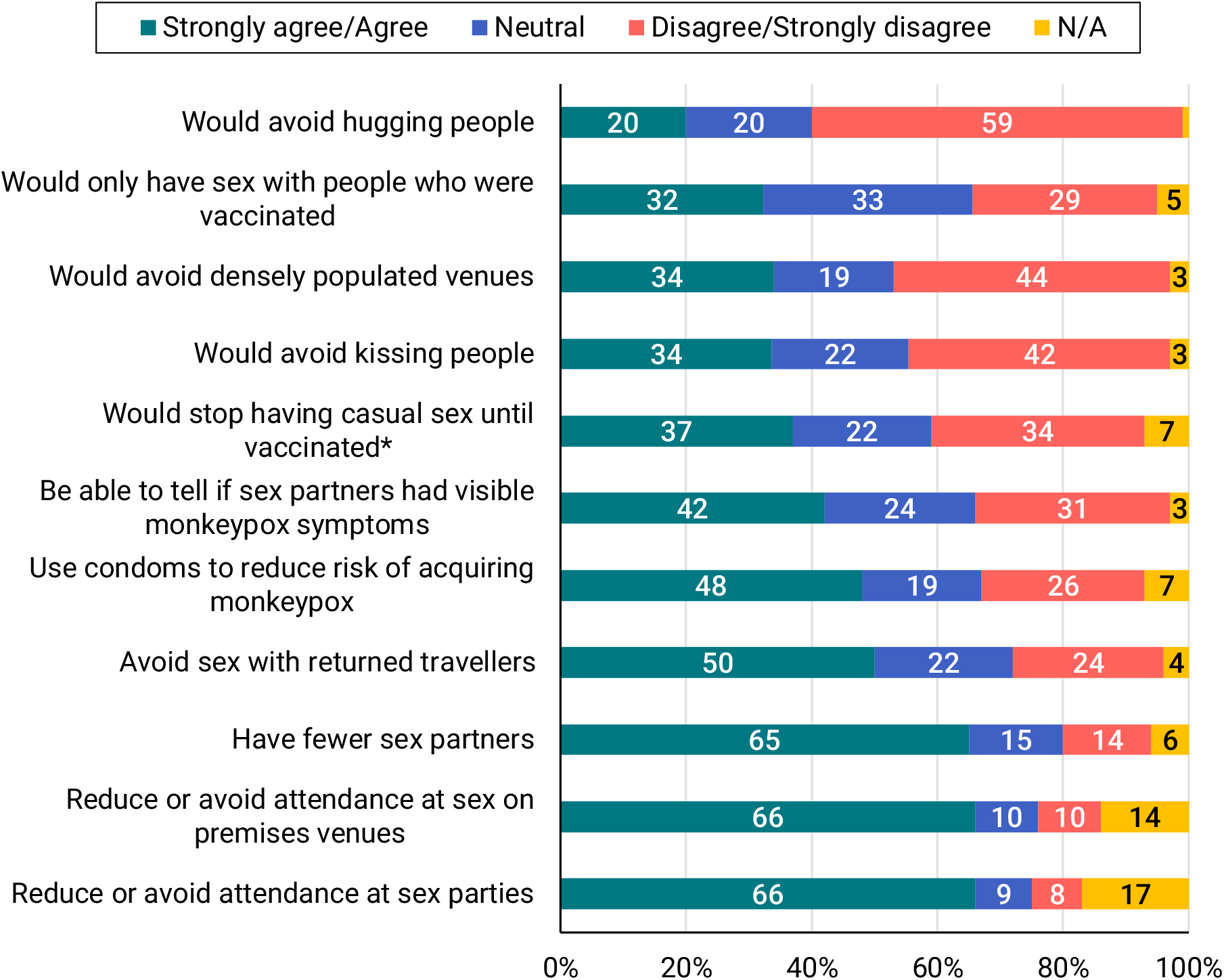
– Endorsement of potential mpox risk-reduction strategies by 2268 participants who had not been diagnosed with mpox. *This item was only shown to the 1733 participants who had not received a smallpox/mpox vaccination or been diagnosed with mpox. All items were presented in random order.

Multivariable analyses showed that concern about mpox and vaccination since May 2022 were independently associated with greater acceptability of these three strategies (see Appendix 2, Table 6 for multivariable results). The least acceptable strategies were not hugging people (445; 19.62%), avoiding densely populated venues (760; 33.51%), and not kissing people (771; 33.99%). Most participants were willing to avoid contact with people if diagnosed with mpox (2110/2268; 93.03%), but fewer participants were confident they could avoid physical contact with people (1660/2268; 73.19%), work from home (1588/2268; 70.02%), or not share communal living spaces such as a bathroom, kitchen or bedroom (1222/1268; 53.88%). More than half of participants were comfortable with contact tracers disclosing a potential mpox diagnosis to casual sex partners (1377/2268; 60.71%; see Appendix 2, Table 7 for further details).

### Willingness to be vaccinated against mpox

Among the 1733 participants who were unvaccinated and had not been diagnosed with mpox, 1457 were willing to be vaccinated (84.07%). Most were willing to receive the vaccine immediately as a precautionary measure (1337/1733; 77.15%); a smaller proportion indicated they would be vaccinated later if more mpox cases were reported in Australia (268/1733; 15.46%). Among the 1486 participants who had at least one male casual sex partner in the previous six months, vaccine acceptability was similar to the broader group (1275/1486, 85.80% vs. 1457/1733, 84.07%). Multivariable analysis showed bisexual participants and those who were unconcerned about mpox were less willing to be vaccinated and participants with greater numbers of recent sexual partners were more willing (see Appendix 2, Table 8 for multivariable results).

## Discussion

This was the first national study of knowledge and attitudes to mpox among GBMSM in Australia. Most participants accurately identified common clinical symptoms (skin lesions and/or rash) and transmission routes (skin-to-skin contact). Early recognition of symptoms is an important component of public health responses to an infectious disease outbreak.

Nearly one quarter of participants reported a history of smallpox or mpox vaccination. As Australia never had universal smallpox vaccination (18, 19) and estimates suggest only 10% of contemporary Australians have been vaccinated against smallpox (20), it is likely that some participants mistakenly believed they had been vaccinated (21). It is unknown whether mistaken beliefs about historical vaccination may deter people from seeking vaccination against mpox, but it may be important for public education campaigns to highlight the limited coverage and protection offered by historical smallpox vaccination (7).

As we anticipated, almost two thirds of participants were willing to modify their sexual practices to reduce their risk of acquiring mpox (e.g., avoiding sex parties, sex-on-premises venues or having fewer sex partners). However, changes to social practices, such as avoiding hugging or kissing, or avoiding popular venues, were rated as less acceptable. It is possible that changes to social behaviour were perceived as less acceptable after COVID-19 restrictions in Australia (‘pandemic fatigue’) (22) and because GBMSM correctly surmised that prolonged skin-to-skin contact represents a greater transmission risk. Previous research has found that GBMSM are unwilling to avoid kissing to reduce the risk of gonorrhoea (23). However, COVID-19 research showed that GBMSM modified their sexual practices to reduce transmission risk, particularly in the absence of vaccine protection (15, 17), gradually increasing levels of sexual activity after they were vaccinated (24). It is difficult for people to sustain behaviour that has a personal or social cost over time (25), so public health messages should consider what behaviour change may be achievable and acceptable for GBMSM during mpox outbreaks, in order for messaging to be effective. Ongoing surveillance of sexual behaviour, vaccine uptake, and behaviour change in response to mpox would be useful.

Lastly, there was very high willingness to receive mpox vaccination as it becomes available. Few factors distinguished between participants who were willing to be vaccinated or not, although bisexual participants and people who were less concerned about mpox were less willing, and people with more sexual partners were more willing. In general, this aligns with community-oriented messaging about mpox vaccination, which encouraged more sexually active people to come forward first, when vaccine supplies were limited. Our results suggest that targeted messaging for bisexual men may be warranted, explaining why vaccination is beneficial, and how vaccination may be accessed safely and discreetly. Such messaging may be particularly important to increase mpox vaccination rates to reduce the risk of future outbreaks, given mpox may circulate globally at low levels for the foreseeable future.

## Limitations

We acknowledge the limitations of the analysis, which included the study’s non-random sample. As there are limited national data on GBMSM and non-binary people, the representativeness of our sample is unknown. However, the cross-sectional sample comprised a large proportion of people who were at potential risk of acquiring mpox (i.e., who reported potential risk behaviours, recent travel and future travel plans). Further, 19 (14.7%) of the 129 people (as of 12 September 2022) who had been diagnosed with mpox in Australia responded to the survey (9). We also oversampled participants from New South Wales and Victoria, where most mpox cases were reported (3). Our findings may therefore reflect the experiences of people with greater interest in mpox or those at risk of acquiring mpox in Australia.

## Data Availability

A deidentified dataset may be available from the authors upon reasonable request, subject to ethical oversight being obtained by bona fide researchers.

## Conclusion

Despite the rapid global emergence of mpox, Australia is well placed to avoid a large-scale local mpox outbreak. This can be achieved through large-scale uptake of mpox vaccination by at-risk people, and targeted education and awareness raising among GBMSM to explain the importance of temporary modifications of sexual practices during outbreaks while vaccination levels increase. Our survey indicates high levels of willingness to receive mpox vaccination, but some populations, namely bisexual men and GBMSM with lower numbers of sexual partners, may benefit from encouragement and support to get vaccinated. Our survey indicates that GBMSM are willing to modify some sexual practices to reduce their risk of mpox, but we caution that such behaviour modification may not be sustainable in the longer term, and hence risk reduction messaging must be carefully timed to achieve maximal impact during periods where there is a risk of transmission. Together, our findings suggest many GBMSM seek to protect their own and others’ sexual health, but some groups may need additional support to access health information, vaccination, and health services. While case numbers are currently low, Australia should seize the opportunity to increase vaccination coverage to reduce the chance of future outbreaks.

## DECLARATIONS

### Author contributions

All authors contributed to the study design, analysis and interpretation of findings. James MacGibbon, Timothy R Broady and Martin Holt oversaw data collection. James MacGibbon drafted the manuscript and conducted the quantitative analyses, supported by Vincent Cornelisse and Martin Holt. All authors reviewed and commented on drafts of the manuscript and agreed with the final version.

## Acknowledgements

In-kind creative services for the study recruitment materials and website were provided by Oskar Westerdal and Vincent Rommelaere. We are grateful to Professor Andrew Grulich and Timmy Lockwood who provided advice on the survey instrument.

## Declaration of funding

The Centre for Social Research in Health and Kirby Institute receive funding from the Australian Government Department of Health and from state/territory health departments. The funding sources did not have any involvement in the collection, analysis and interpretation of data, or in the writing of this manuscript and decision to submit for publication. No pharmaceutical funding was received for this study.

## Conflicts of Interest

Anthony K J Smith is a Joint Editor of *Sexual Health*, but was blinded from the peer-review process for this paper.

## Code availability

Syntax for the database coding and analysis may be available upon request.

## Compliance with ethical standards

The questionnaire and methodology for this study were approved by the Human Research Ethics Committee of UNSW Sydney (HC220484) and endorsed by the community organisation ACON (2022/14). Informed consent was obtained from all individual participants included in the study prior to their participation in the questionnaire.

## Notes

### Competing Interest Statement

The authors have declared no competing interest.

### Author Declarations

The questionnaire and methodology for this study were approved by the Human Research Ethics Committee of the University of New South Wales (UNSW Sydney; HC220484) and endorsed by the community organisation ACON (2022/14). Informed consent was obtained from all individual participants included in the study prior to their participation in the questionnaire.

### Summary of Updates

We made changes to terminology from monkeypox to mpox (disease) and monkeypox virus, consistent with the WHO convention. We also made minor to formatting. The results do not differ.

