## Supplementary materials for "Mpox (monkeypox) knowledge, concern, willingness to change behaviour, and seek vaccination: Results of a national cross-sectional survey"

**APPENDIX 1: Supplementary information about the survey measures**

**Previous smallpox or mpox vaccination**

Participants were provided descriptions of the 3rd-generation, non-replicating Modified Vaccinia Ankara vaccine and the replication-competent 2nd-generation vaccinia vaccine (‘ACAM2000’). Examples of the MVA brand names were provided to aid recognition (‘Jynneous/Imvanex/Imvamune’). Previous vaccination was measured with the item ‘Have you been vaccinated against smallpox or monkeypox? (including at least one vaccine dose)’, with response categories ‘Yes’, ‘No’ and ‘Don’t know’. Participants who selected ‘Yes’ were asked which vaccine they received (MVA; ACAM2000; don’t know), when they were vaccinated (before May 2022; after May 2022), and why they were vaccinated (before potential exposure; after exposure; another reason). Participants who reported receiving the MVA vaccine were asked how many vaccine doses they received and whether they received vaccine dose(s) in Australia, overseas, or both.

**Mpox knowledge and concern**

Mpox knowledge was measured with the item ‘How much do you know about monkeypox virus (MPXV)?’, with response categories as follows: 1=‘I know a lot about it’, 2=‘I know quite a bit about it’, 3=‘I know a fair amount about it’, 4=‘I know a small amount about it’, and 5=‘Nothing: I’ve never heard about it before this survey’. Participants who had heard of mpox (response options 1–4) were then asked ‘How did you learn about monkeypox virus (MPXV)? (select all that apply)’. Response options included items such as ‘Information provided in the media, including TV and social media’ and ‘Searching online/Googling’.

Concern about mpox was measured with the item ‘How concerned are you about getting monkeypox virus (MPXV)?’. Response options ranged from 1=‘Not at all concerned’ to

5=‘Very concerned’. We coded responses 4–5 as concerned about mpox. Responses 1–3 were used as the reference category in the multivariate analyses.

**Mpox symptom and transmission route identification**

Participants who had not been diagnosed with mpox were asked to identify potential mpox symptoms and transmission routes. Recognition of mpox symptoms was measured with the item ‘Can any of the following be symptoms of monkeypox virus (MPXV)?’, with response categories ‘Yes’, ‘No’ and ‘Don’t know’. Participants were asked to select ‘Don’t know’ if they were not sure about symptoms. Twelve potential symptoms were shown in random order, for example, ‘Skin lesions, sores or pustules’, ‘A skin rash’ and ‘Tiredness/Fatigue’. All potential symptoms are shown in Box 1.

Recognition of mpox transmission routes was measured with the item ‘Can monkeypox virus (MPXV) be spread from someone who has the virus through any of the following?’, with response categories ‘Yes’, ‘No’ and ‘Don’t know’. As in the previous item, participants were asked to select ‘Don’t know’ if they were not sure about transmission routes. Eight potential symptoms were shown in random order, for example, ‘Prolonged skin-to-skin contact’, ‘Kissing’ and ‘Sharing drinks’. All potential transmission routes are shown in Box 2.

**Endorsement of behavioural risk reduction strategies**

Participants who had not been diagnosed with mpox were asked how much they agreed with potential behavioural mpox risk reduction strategies: ‘How much do you agree or disagree with the following statements about reducing risk of getting monkeypox virus (MPXV)?’. Response options ranged from 1=‘Strongly disagree’ to 5=‘Strongly agree’ and 6=“N/A”. We coded responses 4–5 as endorsing the strategy. All behavioural risk reduction strategies are shown in Box 3.

**Willingness to be vaccinated against mpox**

Participants who had not received any doses of smallpox or mpox vaccination and had not been diagnosed with mpox were asked how likely they were to accept a vaccine: ‘If a safe and effective vaccine against monkeypox was available to you, how likely are you get vaccinated?’. Response options ranged from 1=‘Very unlikely’ to 5=‘Very likely’. We coded responses 1–3 as hesitant and 4–5 as willing to be vaccinated. Participants were then asked ‘When would you get vaccinated? (tick all that apply)’. Six response options were shown to participants, for example, ‘Right now as a precautionary measure’, ‘If more cases were reported in Australia’, and ‘I wouldn’t get vaccinated’. To aid the interpretation of non-mutually exclusive items, we report frequencies from the second response option (‘If more cases were reported in Australia’) only where that item was chosen exclusively, or participants did not select ‘Right now as a precautionary measure’.

### APPENDIX 2: Supplementary results

**Table 1. Participant characteristics, HIV and STI testing and status, and PrEP use among all participants**

| | *N* = 2287 |
|---|---|
| Age | |
| <30 | 459 (20.07) |
| 30–39 | 666 (29.12) |
| 40–49 | 506 (22.13) |
| 50+ | 656 (28.68) |
| Sexual identity | |
| Gay* | 1894 (82.82) |
| Bisexual/Pansexual | 271 (11.85) |
| Queer/Another term | 122 (5.33) |
| Gender | |
| Male (cis and trans)* | 2189 (95.71) |
| Non-binary/Different identity | 98 (4.29) |
| State or territory | |
| New South Wales | 860 (37.60) |
| Victoria | 760 (33.23) |
| Queensland | 311 (13.60) |
| Other jurisdictions | 356 (15.57) |
| Residential location | |
| Inner metropolitan area of a capital city | 1286 (56.23) |
| Outer metropolitan area of a capital city | 591 (25.84) |
| Other city/regional/rural or remote area | 391 (17.10) |
| Prefer not to say | 19 (0.83) |
| Country of birth | |
| Australia | 1701 (74.38) |
| Elsewhere | 586 (25.62) |
| Education level | |
| High school/Trade certificate | 737 (32.23) |
| University degree | 1550 (67.77) |
| Employment status | |
| Full time | 1647 (72.02) |
| Part time | 274 (11.98) |
| Student/unemployed/other | 366 (16.00) |
| Income level (AUD) | |
| Less than $40,000 | 329 (14.39) |
| $40,000−$79,999 | 485 (21.21) |
| $80,000−$120,000 | 651 (28.47) |
| More than $120,000 | 663 (28.99) |
| Prefer not to say | 159 (6.95) |
| HIV status | |
| Untested/Unknown status | 164 (7.17) |
| HIV negative | 1944 (85.00) |
| HIV positive | 179 (7.83) |
| Last tested for HIV | |
| ≤12 months | 1634 (71.45) |
| >12 months | 508 (22.21) |
| Never tested for HIV | 145 (6.34) |
| Last tested for sexually transmissible infections (STIs) | |
| ≤12 months | 1616 (70.66) |
| >12 months | 450 (19.68) |
| Never tested for STIs | 221 (9.66) |
| STI diagnosis in the last 12 months | 576 (25.19) |
| Ever used PrEP | 1241 (54.26) |
| Currently using PrEP | 914 (39.97) |

*Note*. Data are *n*(%). *1881 were cisgender gay men (82.25%); 27 participants were transgender male (1.18%).

**Table 2. Previous vaccination against smallpox or mpox among all participants**

| | $N = 2287$ |
|---|---|
| Ever received a smallpox or mpox vaccine | |
| Yes | 541 (23.66) |
| No | 1412 (61.74) |
| Don't know | 334 (14.60) |
| **(Vaccinated only)** | **(*n* = 541)** |
| Vaccination type | |
| Modified vaccinia Ankara (MVA) vaccine | 325 (60.07) |
| Smallpox vaccinia (live) vaccine | 28 (5.18) |
| Don't know | 188 (34.75) |
| Vaccination period | |
| Before May 2022 | 194 (35.86) |
| After May 2022 | 347 (64.14) |
| **(Vaccinated since May 2022 only)** | **(*n* = 347)** |
| Reason for vaccination | |
| After potential exposure to mpox | 15 (4.32) |
| Before potential exposure to mpox | 283 (81.56) |
| Another reason | 42 (12.10) |
| Prefer not to say | 7 (2.02) |
| **(Vaccinated with MVA vaccine only)** | **(*n* = 325)** |
| Number of vaccine doses | |
| One dose | 320 (98.46) |
| Two doses | 3 (0.92) |
| Don't know | 2 (0.62) |

*Note*. Data are *n*(%).

**Table 3. Potential mpox risk factors among all participants by mpox vaccination status**

| | Total $N = 2287$ | Unvaccinated $n = 1942$ | Vaccinated $n = 345$ | $X^2$ | *p* value |
|---|---|---|---|---|---|
| Recent overseas travel | 706 (30.87) | 551 (28.37) | 155 (44.93) | 37.62 | <.001 |
| Planned overseas travel (next 6 months) | 1075 (47.00) | 855 (44.03) | 220 (63.77) | 45.83 | <.001 |
| No. of male sexual partners | | | | 97.37 | <.001 |
| None | 293 (12.81) | 280 (14.42) | 13 (3.77) | | |
| 1–10 | 1395 (61.00) | 1220 (62.82) | 175 (50.72) | | |
| 11–20 | 283 (12.37) | 220 (11.33) | 63 (18.26) | | |
| 20+ | 316 (13.82) | 222 (11.43) | 94 (27.25) | | |
| Sex with casual male partners | | | | 78.47 | <.001 |
| No casual partners/no anal sex | 773 (33.80) | 720 (37.08) | 53 (15.36) | | |
| Consistent condom use | 237 (10.36) | 212 (10.92) | 25 (7.25) | | |
| Any condomless sex | 1277 (55.84) | 1010 (52.01) | 267 (77.39) | | |
| Had group sex | 804 (35.16) | 621 (31.98) | 183 (53.04) | 57.04 | <.001 |
| Had an anonymous sex | 1248 (54.57) | 1007 (51.85) | 241 (69.86) | 38.29 | <.001 |
| Visited a sex on premises venue | 731 (31.96) | 549 (28.27) | 182 (52.75) | 80.75 | <.001 |
| Visited a beat/cruising area | 498 (21.78) | 397 (20.44) | 101 (29.28) | 13.41 | <.001 |
| Been to a private sex party | 185 (8.09) | 125 (6.44) | 60 (17.39) | 47.29 | <.001 |
| Used drugs for sex | 244 (10.67) | 184 (9.47) | 60 (17.39) | 19.26 | <.001 |

*Note*. Data are *n*(%). All recall periods were 6 months preceding the survey. Comparisons were made using Pearson's chi-squared tests. The vaccinated comparison group includes only people who were vaccinated against mpox since May 2022.

**Table 4. Mpox knowledge and concern among all participants excluding those diagnosed with mpox, by mpox vaccination status**

| | Total<br>$N$ = 2268 | Unvaccinated<br>$n$ = 1928 | Vaccinated<br>$n$ = 340 | $X^2$ | $p$ value |
|---|---|---|---|---|---|
| Self-assessed mpox knowledge | | | | 90.49 | <.001 |
| Never heard about it | 32 (1.41) | 32 (1.66) | 0 (0) | | |
| Knew a small amount | 1065 (46.96) | 974 (50.52) | 91 (26.76) | | |
| Knew a fair amount | 695 (30.64) | 569 (29.51) | 126 (37.06) | | |
| Knew quite a bit | 356 (15.70) | 267 (13.85) | 89 (26.18) | | |
| Knew a lot | 120 (5.29) | 86 (4.46) | 34 (10.00) | | |
| Knows people diagnosed with mpox | 323 (14.24) | 226 (11.72) | 97 (28.53) | 66.85 | <.001 |
| Concern about acquiring mpox | | | | 37.24 | <.001 |
| Not at all/Not concerned | 382 (16.84) | 357 (18.52) | 25 (7.35) | | |
| Neither | 425 (18.74) | 376 (19.50) | 49 (14.41) | | |
| Concerned/Very concerned | 1461 (64.42) | 1195 (61.98) | 266 (78.24) | | |
| Perceived likelihood of acquiring mpox | | | | 12.65 | .002 |
| Very unlikely/Unlikely | 984 (43.39) | 864 (44.81) | 120 (35.29) | | |
| Neither | 819 (36.11) | 670 (34.75) | 149 (43.82) | | |
| Likely/Very likely | 465 (20.50) | 394 (20.44) | 71 (20.88) | | |

*Note*. Data are *n*(%). Comparisons were made using Pearson's chi-squared tests. The vaccinated comparison group only includes people who were vaccinated against mpox since May 2022.

**Table 5. Mpox symptom and transmission route identification among 2268 participants who had not been diagnosed with mpox, by recent male sexual partner numbers**

| | Total<br>$N$ = 2268 | Up to 10 partners<br>$n$ = 1928 | More than 10 partners<br>$n$ = 340 | $X^2$ | $p$ value |
|---|---|---|---|---|---|
| Potential symptoms identified | | | | | |
| Skin lesions | 2087 (92.02) | 1756 (91.08) | 331 (97.35) | 15.49 | <.001 |
| Skin rash | 1977 (87.17) | 1655 (85.84) | 322 (94.71) | 20.31 | <.001 |
| Fever | 1647 (72.62) | 1347 (69.87) | 300 (88.24) | 49.05 | <.001 |
| Tiredness/fatigue | 1576 (69.49) | 1291 (66.96) | 285 (83.82) | 38.76 | <.001 |
| Swollen glands | 1509 (66.53) | 1238 (64.21) | 271 (79.71) | 31.16 | <.001 |
| Muscle aches | 1386 (61.11) | 1117 (57.94) | 269 (79.12) | 54.57 | <.001 |
| Headaches | 1335 (58.86) | 1088 (56.43) | 247 (72.65) | 31.89 | <.001 |
| Chills | 1218 (53.70) | 974 (50.52) | 244 (71.76) | 52.47 | <.001 |
| Anal pain and/or bleeding | 887 (39.11) | 688 (35.68) | 199 (58.53) | 63.34 | <.001 |
| Potential transmission routes identified | | | | | |
| Prolonged skin-to-skin contact | 2124 (93.65) | 1786 (92.63) | 338 (99.41) | 22.32 | <.001 |
| Brief skin-to-skin contact | 1860 (82.01) | 1570 (81.43) | 290 (85.29) | 2.92 | .09 |
| Bodily fluids | 1692 (74.60) | 1427 (74.01) | 265 (77.94) | 2.35 | .13 |
| Kissing | 1664 (73.37) | 1390 (72.10) | 274 (80.59) | 10.67 | .001 |
| Sharing a bed | 1633 (72.00) | 1343 (69.66) | 290 (85.29) | 35.05 | <.001 |
| Contact with clothes of an infected person | 1357 (59.83) | 1111 (57.62) | 246 (72.35) | 26.09 | <.001 |
| Coughing/sneezing (breathing aerosols) | 983 (43.34) | 803 (41.65) | 180 (52.94) | 15.01 | <.001 |
| Sharing drinks | 824 (36.33) | 694 (36.00) | 130 (38.24) | 0.63 | .43 |

*Note*. Data are *n*(%). Proportions represent affirmative ('Yes') responses. Comparisons were made using Pearson's chi-squared tests. Dichotomised partner numbers relate to male sexual partners in the six months preceding the survey.

**Table 6. Factors affecting acceptability of common social and behavioural changes in response to mpox among 2268 participants who had not been diagnosed with mpox**

| | Does not support the strategy* | Supports the strategy | OR (95% CI) | *p* value | aOR (95% CI) | *p* value |
|---|---|---|---|---|---|---|
| **Reduce or avoid attendance at sex parties** | ***n* = 774 [34.13]** | ***n* = 1494 [65.87]** | | | | |
| Concerned about mpox | 427 (55.17) | 1034 (69.21) | 1.83 (1.53–2.19) | <.001 | 1.71 (1.43–2.05) | <.001 |
| Current PrEP user | 309 (39.92) | 593 (39.69) | 0.99 (0.83–1.18) | .92 | – | |
| Diagnosed with an STI (last 12mths) | 157 (20.28) | 411 (27.51) | 1.49 (1.21–1.84) | <.001 | 1.27 (1.02–1.58) | .03 |
| Vaccinated for MPX since May 2022 | 85 (10.98) | 255 (17.07) | 1.67 (1.28–2.17) | <.001 | 1.45 (1.11–1.90) | .007 |
| >10 recent male partners (last 6mths) | 197 (25.45) | 389 (26.04) | 1.03 (0.85–1.26) | .76 | – | |
| **Reduce or avoid attendance at sex on premises venues** | ***n* = 762 [33.60]** | ***n* = 1506 [66.40]** | | | | |
| Concerned about mpox | 417 (54.72) | 1044 (69.32) | 1.87 (1.56–2.24) | <.001 | 1.78 (1.48–2.13) | <.001 |
| Current PrEP user | 310 (40.68) | 592 (39.31) | 0.94 (0.79–1.13) | .53 | – | |
| Diagnosed with an STI (last 12mths) | 162 (21.26) | 406 (26.96) | 1.37 (1.11–1.68) | .003 | 1.16 (0.93–1.43) | .19 |
| Vaccinated for MPX since May 2022 | 85 (11.15) | 255 (16.93) | 1.62 (1.25–2.11) | <.001 | 1.43 (1.09–1.88) | .009 |
| >10 recent male partners (last 6mths) | 212 (27.82) | 374 (24.83) | 0.86 (0.70–1.04) | .12 | – | |
| **Have fewer sex partners** | ***n* = 802 [35.36]** | ***n* = 1466 [64.64]** | | | | |
| Concerned about mpox | 437 (54.49) | 1024 (69.85) | 1.94 (1.62–2.31) | <.001 | 2.11 (1.75–2.54) | <.001 |
| Current PrEP user | 332 (41.40) | 570 (38.88) | 0.90 (0.76–1.07) | .24 | – | |
| Diagnosed with an STI (last 12mths) | 184 (22.94) | 384 (26.19) | 1.19 (0.97–1.46) | .09 | – | |
| Vaccinated for MPX since May 2022 | 88 (10.97) | 252 (17.19) | 1.68 (1.30–2.18) | <.001 | 1.78 (1.36–2.34) | <.001 |
| >10 recent male partners (last 6mths) | 247 (30.80) | 339 (23.12) | 0.68 (0.56–0.82) | <.001 | 0.52 (0.42–0.63) | <.001 |

*Note.* Data are n(%), and unadjusted odds ratios and adjusted odds ratios with 95% confidence intervals derived from logistic regression. The ‘Does not support the strategy’ dependent variable category includes ‘Strongly disagree’, ‘Disagree’, ‘Neither disagree nor agree’ and ‘Not applicable’ responses. The ‘Supports the strategy’ category includes ‘Agree’ and ‘Strongly agree’. Square brackets enclose row proportions for each strategy. Strategies are displayed in the table by level of support in descending order, however survey items were randomised to participants to reduce response order bias.

The reference categories for independent variables were: 0 = ‘Not concerned about monkeypox’; 0 = ‘Not a current PrEP user’; 0 = ‘Had not been diagnosed with an STI in the 12 months preceding the survey’; 0 = ‘Had not received a smallpox or monkeypox vaccine since May 2022’; 0 = ‘Had up to 10 male sexual partners in the six months preceding the survey’.

**Table 7. Comfort with contact tracing, and willingness and perceived ability to comply with potential isolation requirements among 2268 participants who had not been diagnosed with mpox**

| | *N* = 2268 |
|---|---|
| Comfortable with contact tracers disclosing a potential MPX diagnosis to: | |
| Regular sex partners | 1635 (72.09) |
| Casual sex partners | 1431 (63.10) |
| Friends | 1377 (60.71) |
| Family | 962 (42.42) |
| Workplace/colleagues | 831 (36.64) |
| Willingness to avoid sexual contact with partners during isolation | 2191 (96.60) |
| Willingness to take antiviral drugs to reduce mpox severity* | 2174 (95.86) |
| Willingness to avoid contact with people during isolation | 2110 (93.03) |
| Willingness to use condoms for two months after infection | 1832 (80.78) |
| Confident in ability to wear a mask in public if diagnosed | 2100 (92.59) |
| Confident in ability to avoid physical contact if diagnosed | 1660 (73.19) |
| Confident in ability to work from home if diagnosed | 1588 (70.02) |
| Confident in ability to avoid contact with pets or animals if diagnosed | 1419 (62.57) |
| Confident in ability to not share communal living areas | 1222 (53.88) |

*Note*. Data are *n*(%). Details of potential side effects and dosing regimens for antiviral medications were not provided in the survey and willingness should therefore be interpreted with caution. Items were measured on 5 and 6–point Likert-type scales and were dichotomised based on the following: ‘Comfort’ = ‘Somewhat comfortable’ and ‘Very comfortable’; Willing = ‘Willing’ and ‘Very willing’; and ‘Confident’ = ‘Confident’ and ‘Very confident’. The contact tracing items included a ‘Not applicable’ (N/A) response option, responses to which have been coded as ‘Neutral/Don’t know’. Excluding the N/A responses from the denominators increases the levels of comfort in contact tracing with regular and casual sex partners (1635/2139, 76.44%; and 691/2122, 67.44%, respectively).

**Table 8. Factors affecting willingness to receive a mpox vaccine among 1486 participants who had not been diagnosed with mpox, had not received a smallpox or mpox vaccine and had at least one recent casual sex partner**

| | Total<br>*N* = 1486 | Unwilling to be vaccinated<br>*n* = 211 | Willing to be vaccinated<br>*n* = 1275 | OR (95% CI) | *p* value | aOR (95% CI) | *p* value |
|---|---|---|---|---|---|---|---|
| Age | | | | | | | |
| <30 | 339 (22.81) | 43 (20.38) | 296 (23.22) | Ref | | | |
| 30–39 | 437 (29.41) | 57 (27.01) | 380 (29.80) | 0.97 (0.63–1.48) | .882 | | |
| 40–49 | 334 (22.48) | 50 (23.70) | 284 (22.27) | 0.83 (0.53–1.28) | .391 | | |
| 50+ | 376 (25.30) | 61 (28.91) | 315 (24.71) | 0.75 (0.49–1.14) | .181 | | |
| Sexual identity | | | | | | | |
| Gay | 1247 (83.92) | 165 (78.20) | 1082 (84.86) | Ref | | Ref | |
| Bisexual/Pansexual | 171 (11.51) | 39 (18.48) | 132 (10.35) | 0.52 (0.35–0.76) | .001 | 0.63 (0.41–0.96) | **.032** |
| Queer/Another term | 68 (4.58) | 7 (3.32) | 61 (4.78) | 1.33 (0.60–2.95) | .486 | 1.18 (0.49–2.84) | .717 |
| State or territory | | | | | | | |
| New South Wales | 538 (36.20) | 78 (36.97) | 460 (36.08) | Ref | | | |
| Victoria | 482 (32.44) | 63 (29.86) | 419 (32.86) | 1.13 (0.79–1.61) | .51 | | |
| Queensland | 224 (15.07) | 38 (18.01) | 186 (14.59) | 0.83 (0.54–1.27) | .388 | | |
| Other states/territories | 242 (16.29) | 32 (15.17) | 210 (16.47) | 1.11 (0.71–1.73) | .636 | | |
| Residential location | | | | | | | |
| Inner metro | 811 (54.58) | 100 (47.39) | 711 (55.76) | Ref | | Ref | |
| Outer metro | 401 (26.99) | 59 (27.96) | 342 (26.82) | 0.82 (0.58–1.15) | .248 | 0.97 (0.66–1.41) | .858 |
| Regional and remote | 256 (17.23) | 48 (22.75) | 208 (16.31) | 0.61 (0.42–0.89) | .01 | 0.82 (0.54–1.27) | .377 |
| No response | 18 (1.21) | 4 (1.90) | 14 (1.10) | 0.49 (0.16–1.53) | .219 | 0.48 (0.17–1.36) | .165 |
| Country of birth | | | | | | | |
| Australia | 1139 (76.65) | 161 (76.30) | 978 (76.71) | Ref | | | |
| Elsewhere | 347 (23.35) | 50 (23.70) | 297 (23.29) | 0.98 (0.69–1.38) | .898 | | |
| Education level | | | | | | | |
| High school | 234 (15.75) | 47 (22.27) | 187 (14.67) | Ref | | Ref | |
| Trade certificate | 243 (16.35) | 37 (17.54) | 206 (16.16) | 1.40 (0.87–2.25) | .165 | 1.35 (0.82–2.22) | .236 |
| University degree | 1009 (67.90) | 127 (60.19) | 882 (69.18) | 1.75 (1.21–2.53) | .003 | 1.32 (0.87–1.98) | .188 |
| Employment status | | | | | | | |
| Full time | 1101 (74.09) | 160 (75.83) | 941 (73.80) | Ref | | | |
| Part time | 168 (11.31) | 21 (9.95) | 147 (11.53) | 1.19 (0.73–1.94) | .483 | | |
| Student/Unemployed/Other | 217 (14.60) | 30 (14.22) | 187 (14.67) | 1.06 (0.70–1.61) | .786 | | |

**CONT'D**

| | Total<br>$N = 1486$ | Unwilling to be vaccinated<br>$n = 211$ | Willing to be vaccinated<br>$n = 1275$ | OR (95% CI) | *p* value | aOR (95% CI) | *p* value |
|---|---|---|---|---|---|---|---|
| Income level (AUD) | | | | | | | |
| Less than $40,000 | 207 (13.93) | 27 (12.80) | 180 (14.12) | Ref | | | |
| $40,000−$79,999 | 324 (21.80) | 48 (22.75) | 276 (21.65) | 0.86 (0.52–1.43) | .568 | | |
| $80,000−$120,000 | 427 (28.73) | 61 (28.91) | 366 (28.71) | 0.90 (0.55–1.46) | .671 | | |
| More than $120,000 | 425 (28.60) | 56 (26.54) | 369 (28.94) | 0.99 (0.60–1.62) | .963 | | |
| Prefer not to say | 103 (6.93) | 19 (9.00) | 84 (6.59) | 0.66 (0.35–1.26) | .21 | | |
| Concern about acquiring mpox | | | | | | | |
| Not at all/Not concerned | 203 (13.66) | 76 (36.02) | 127 (9.96) | Ref | | Ref | |
| Neither | 275 (18.51) | 44 (20.85) | 231 (18.12) | 3.14 (2.04–4.83) | <.001 | 3.26 (2.10–5.06) | **<.001** |
| Concerned/Very concerned | 1008 (67.83) | 91 (43.13) | 917 (71.92) | 6.03 (4.22–8.61) | <.001 | 5.18 (3.36–7.99) | **<.001** |
| Perceived likelihood of acquiring mpox | | | | | | | |
| Very unlikely/Unlikely | 591 (39.77) | 114 (54.03) | 477 (37.41) | Ref | | Ref | |
| Neither | 550 (37.01) | 69 (32.70) | 481 (37.73) | 1.67 (1.20–2.31) | .002 | 0.88 (0.60–1.28) | .492 |
| Likely/Very likely | 345 (23.22) | 28 (13.27) | 317 (24.86) | 2.71 (1.75–4.19) | <.001 | 1.09 (0.66–1.82) | .734 |
| Self-assessed health status | | | | | | | |
| All else | 614 (41.32) | 83 (39.34) | 531 (41.65) | Ref | | | |
| Very good/Excellent | 872 (58.68) | 128 (60.66) | 744 (58.35) | 0.91 (0.67–1.22) | .528 | | |
| Knows people diagnosed with mpox | 192 (12.92) | 21 (9.95) | 171 (13.41) | 1.40 (0.87–2.26) | 0.167 | | |
| HIV status | | | | | | | |
| HIV negative | 1303 (87.69) | 175 (82.94) | 1128 (88.47) | Ref | | Ref | |
| HIV positive | 93 (6.26) | 14 (6.64) | 79 (6.20) | 0.88 (0.49–1.58) | 0.659 | 0.86 (0.45–1.63) | 0.635 |
| Untested/unknown | 90 (6.06) | 22 (10.43) | 68 (5.33) | 0.48 (0.29–0.80) | 0.004 | 0.69 (0.40–1.18) | 0.173 |
| Currently using PrEP | 601 (40.44) | 61 (28.91) | 540 (42.35) | 1.81 (1.31–2.48) | <.001 | 1.17 (0.78–1.76) | 0.435 |
| No. of male sexual partners | | | | | | | |
| 1–10 | 1092 (73.49) | 178 (84.36) | 914 (71.69) | Ref | | Ref | |
| 11–20 | 198 (13.32) | 15 (7.11) | 183 (14.35) | 2.38 (1.37–4.12) | .002 | 1.85 (1.02–3.37) | **.044** |
| More than 20 | 196 (13.19) | 18 (8.53) | 178 (13.96) | 1.93 (1.16–3.21) | .012 | 1.57 (0.85–2.88) | .146 |

**CONT'D**

| | Total<br><br>$N$ = 1486 | Unwilling to be vaccinated<br>$n$ = 211 | Willing to be vaccinated<br>$n$ = 1275 | OR (95% CI) | $p$ value | aOR (95% CI) | $p$ value |
|---|---|---|---|---|---|---|---|
| Sex with casual male partners | | | | | | | |
| No casual partners/no anal sex | 387 (26.04) | 75 (35.55) | 312 (24.47) | Ref | | | |
| Consistent condom use | 195 (13.12) | 22 (10.43) | 173 (13.57) | 1.89 (1.13–3.15) | .014 | 1.33 (0.76–2.33) | .322 |
| Any condomless sex | 904 (60.83) | 114 (54.03) | 790 (61.96) | 1.67 (1.21–2.29) | .002 | 1.01 (0.66–1.54) | .961 |
| STI diagnosis in the last 12 months | 364 (24.50) | 37 (17.54) | 327 (25.65) | 1.62 (1.11–2.36) | .012 | 1.11 (0.74–1.67) | .61 |
| Had group sex | 559 (37.62) | 66 (31.28) | 493 (38.67) | 1.39 (1.01–1.89) | .041 | 0.97 (0.64–1.46) | .878 |
| Had an anonymous sex | 909 (61.17) | 113 (53.55) | 796 (62.43) | 1.44 (1.07–1.93) | .015 | 0.94 (0.63–1.41) | .774 |
| Visited an SOPV | 490 (32.97) | 53 (25.12) | 437 (34.27) | 1.55 (1.12–2.17) | .009 | 1.02 (0.68–1.54) | .908 |
| Visited a beat/cruising area | 366 (24.63) | 43 (20.38) | 323 (25.33) | 1.33 (0.93–1.90) | .123 | | |
| Been to a private sex party | 104 (7.00) | 15 (7.11) | 89 (6.98) | 0.98 (0.56–1.73) | .946 | | |
| Used drugs for sex | 157 (10.57) | 20 (9.48) | 137 (10.75) | 1.15 (0.70–1.88) | .58 | | |
| Overseas travel plans (next 6 months) | 679 (45.69) | 91 (43.13) | 588 (46.12) | 1.13 (0.84–1.51) | .42 | | |

*Note.* Data are n(%), and unadjusted odds ratios and adjusted odds ratios derived from logistic regression. All recall periods were 6 months preceding the survey. Bolded values represent significance at $p$<.05 for ease of reference. The dependent variable survey item was as follows: ‘If a safe and effective vaccine against monkeypox was available to you, how likely are you get vaccinated? Assume for this question that the vaccine being offered would be a modified vaccinia Ankara (MVA) vaccine’. The ‘Unwilling to be vaccinated’ dependent variable category includes ‘Very unlikely, ‘Unlikely’, and ‘Neutral/Don’t know’ responses. The ‘Willing to be vaccinated’ category includes ‘Somewhat likely’ and ‘Very likely’.
